# Predicting SARS-CoV-2 infections for children and youth with single symptom screening

**DOI:** 10.1101/2021.08.19.21262310

**Authors:** Richard J. Webster, Deepti Reddy, Mary-Ann Harrison, Ken J. Farion, Jacqueline Wilmore, Michelle Foote, Nisha Thampi

**Affiliations:** Children’s Hospital of Eastern Ontario Research Institute, Ottawa, Ontario, Canada; Department of Pediatrics, Faculty of Medicine, University of Ottawa, Children’s Hospital of Eastern Ontario, Ottawa; Ottawa Public Health, Ottawa; Department of Family Medicine, Queen’s University, Kingston, Ontario, Canada

## Abstract

Symptom-based SARS-CoV-2 screening and testing decisions in children have important implications on daycare and school exclusion policies. Single symptoms account for a substantial volume of testing and disruption to in-person learning and childcare, yet their predictive value is unclear, given the clinical overlap with other circulating respiratory viruses and non-infectious etiologies. We aimed to determine the relative frequency and predictive value of single symptoms for paediatric SARS-CoV-2 infections from an Ottawa COVID-19 assessment centre from October 2020 through April 2021.

Overall, 46.3% (n=10,688) of pediatric encounters were for single symptoms, and 2.7% of these tested positive. The most common presenting single symptoms were rhinorrhea (31.8%), cough (17.4%) and fever (14.0%). Among children with high-risk exposures children in each age group, the following single symptoms had a higher proportion of positive SARS-CoV-2 cases compared to no symptoms; fever and fatigue (0-4 years); fever, cough, headache, and rhinorrhea (5-12 years); fever, loss of taste or smell, headache, rhinorrhea, sore throat, and cough (13-17 years). There was no evidence that the single symptom of either rhinorrhea or cough predicted SARS-CoV-2 infections among 0-4 year olds, despite accounting for a large volume (61.1%) of single symptom presentations in the absence of high-risk exposures.

Symptom-based screening needs to be responsive to changes in evidence and local factors, including the expected resurgence of other respiratory viruses following relaxation of social distancing/masking, to reduce infection-related risks in schools and daycare settings.

## Introduction

Enhanced health and safety measures have helped to reduce the spread of severe acute respiratory syndrome coronavirus 2 (SARS-CoV-2) among students, parents and staff in schools.^1-8^ A key strategy has been symptom-based screening, exclusion from school, and testing for SARS-CoV-2.^6, 9^ However, more than one-third of children who test positive for SARS-CoV-2 report no symptoms, and clinical manifestations of coronavirus disease 2019 (COVID-19) in children are indistinguishable from other respiratory viral infections, particularly rhinorrhea/congestion and sore throat/dysphagia.^5, 10^ Further, other respiratory viruses are likely to increase as community-based public health measures are relaxed.^11^

Many symptom-based screening policies in schools operating during the pandemic followed a precautionary principle of testing and excluding students with any symptom(s) possibly associated with SARS-CoV-2 infection, even if those symptoms were not conventionally associated with infection transmission risks (e.g. headache, abdominal pain, anosmia). There are costs to the healthcare system to perform unnecessary testing in addition to implications of keeping children out of schools, including learning disruption and caregiver productivity losses.^12, 13^ As circulating non-SARS-CoV-2 respiratory pathogens increase, symptom-based testing for SARS-CoV-2 will determine which students can return when symptoms improve, and which students must continue to isolate due to a positive or unknown COVID-19 status.^14^ Given that children under 12 years of age remain vulnerable to infection as a vaccine-ineligible population, and that they may present initially with single symptoms, knowing which symptoms are more likely to be associated with SARS-CoV-2 infection may help to direct screening, exclusion and testing strategies among school-aged children and youth.

Research into pediatric evidence-based symptom screening is important.^15, 16^ Currently there is an evidence gap for symptom-based screening, with no studies evaluating the predictive value of single symptoms. For example, children and youth may be dismissed from the classroom with only rhinorrhea, but it remains unclear how well any single symptom predicts SARS-CoV-2 infection. Decision makers need these data to balance detection of COVID-19 cases while minimising the disturbance to families.^17^

The objectives of this study are to i) describe the relative frequency of single symptoms, and ii) determine the predictive value of single symptom presentations for SARS-CoV-2 infections among children/youth.

## Methods

### Patient population

This was a retrospective cohort study of children between 0 and 17 years old (daycare-aged [0-4 years], primary school-aged [5-12 years]; secondary school-aged [13-17 years]) who were tested at a pediatric hospital-run COVID-19 assessment centre in Ottawa, Canada, between October 1, 2020, and April 25, 2021. This centre completes more than three-quarters of all pediatric SARS-CoV-2 tests in this city of one million residents. Data were captured in an electronic health record, and the pediatric hospital’s Research Ethics Board approved this secondary use of health administrative data.

Inclusion criteria for this study were patients with a nasopharyngeal or oral-nasal SARS-CoV-2 PCR test who presented with a single symptom and those who were asymptomatic following a high-risk exposure (HRE). Exclusion criteria were non-Ontario residents, children presenting with more than one symptom, a previous confirmed SARS-CoV-2 infection, and encounters with incomplete data.

### Symptoms

In this study, single symptoms were classified according to the Ontario Ministry of Health’s list of symptoms to be screened.^18^ Only symptoms with more than five encounters were included in this study for privacy reasons. Each record represents a separate screening test encounter, with multiple records per child possible. Symptoms were self- or proxy-reported. While our study selection criteria were consistent, the Ontario provincial criteria for testing changed during the course of the study. Until February 21, 2021, children with a single mild symptom (e.g., rhinorrhea/congestion, sore throat) were advised to present for testing only if symptoms persisted >24 hours and/or new symptoms emerged.^19^ As the incidence of SARS-CoV-2 in Ontario increased, stricter screening measures were applied, so that from February 22, 2021, testing guidelines allowed for early presentations with any one symptom. Patients with a HRE were either directed for testing by the local public health unit through the course of case investigations, or self-identified and presented for testing.

### Statistical methods

Within each age group, data were stratified into two cohorts: (1) all children presenting with a single symptom; and (2) those with a HRE who presented with either a single symptom or no symptoms. Provincial regulations generally prevented asymptomatic non-HRE individuals from being tested at COVID-19 assessment centres, and thus the HRE cohort enabled determination of additive linear increases in risk across single symptoms, and may have reduced selection bias across symptom severity (*sensu* Berkson’s paradox).

PPV was reported with 95% CI (derived using the Wilson’s score method).^20^ In the HRE group, to test for differences between the PPV of single and no symptoms, a two proportion Z-test with Holm’s false-discovery correction was used. Likelihood ratios were bootstrapped using the R function bootLR::BayesianLR.test^21^ and their 95% CIs presented. All analyses were performed using R statistical software version 3.5.2.^22^

## Results

During the 29-week study period, there were 54,615 patient encounters, of which 73.3% (40,060/54,615) involved symptoms. Among encounters, 23,080 had single symptoms or were asymptomatic (Fig 1). Almost all patients in the study period had a nasopharyngeal swab; 0.5% had an oral-nasal swab. Among encounters for single-symptom testing, 4,851 were in children aged 0-4 years, 4,763 in children aged 5-12 years, and 1,074 in children aged 13-17 years. Cohort demographic and their clinical presentations are shown in Table 1. A time-series of SARS-CoV-2 rates in Ottawa is provided in Supplemental Fig 1, with trends consistent across age groups.

**Table 1.** Demographics and clinical characteristics of patients presenting with a single symptom at a COVID-19 assessment centre.

|  | **Total**<br>N = 10688 | **Daycare**<br>**Aged**<br>**(0-4 years)**<br>n = 4851 | **Primary**<br>**School Aged**<br>**(5-12 years)**<br>n = 4763 | **Secondary**<br>**School Aged**<br>**(13-17 years)**<br>n = 1074 |
| --- | --- | --- | --- | --- |
| **Age** |  |  |  |  |
| Mean ± SD | 6.0 ± 4.3 | 2.3 ± 1.2 | 7.8 ± 2.2 | 14.8 ± 1.5 |
| Median (IQR) | 5.0 (2.0, 9.0) | 2.0 (1.0, 3.0) | 8.0 (6.0, 10.0) | 15.0 (13.0, 16.0) |
| **Gender, n (%)** |  |  |  |  |
| Male | 5768 (54.0) | 2594 (53.5) | 2550 (53.5) | 624 (58.1) |
| **SARS-CoV-2 Test Result, n (%)** |  |  |  |  |
| Positive | 289 (2.7) | 80 (1.6) | 137 (2.9) | 72 (6.7) |
| **Single Symptom, n (%)** |  |  |  |  |
| Rhinorrhea/congestion | 3396 (31.8) | 2098 (43.2) | 1158 (24.3) | 140 (13.0) |
| Cough | 1860 (17.4) | 866 (17.9) | 822 (17.3) | 172 (16.0) |
| Fever | 1500 (14.0) | 1126 (23.2) | 313 (6.6) | 61 (5.7) |
| Sore throat/difficulty swallowing | 1356 (12.7) | 108 (2.2) | 928 (19.5) | 320 (29.8) |
| Nausea/vomiting | 936 (8.8) | 263 (5.4) | 541 (11.4) | 132 (12.3) |
| Headache | 821 (7.7) | 21 (0.4) | 646 (13.6) | 154 (14.3) |
| Diarrhea | 492 (4.6) | 282 (5.8) | 181 (3.8) | 29 (2.7) |
| Fatigue | 131 (1.2) | 28 (0.6) | 69 (1.4) | 34 (3.2) |
| Sneezing | 121 (1.1) | 55 (1.1) | 61 (1.3) | 5 (0.5) |
| Shortness of breath | 32 (0.3) | 2 (0.0) | 18 (0.4) | 12 (1.1) |
| Muscle/body ache | 27 (0.3) | 2 (0.0) | 19 (0.4) | 6 (0.6) |
| Loss of taste or smell | 16 (0.1) | 0 (0.0) | 7 (0.1) | 9 (0.8) |
| **High Risk Exposure, n (%)** |  |  |  |  |
| Yes | 1140 (10.7) | 326 (6.7) | 579 (12.2) | 235 (21.9) |

<sup>§</sup> In the daycare-aged HRE cohort, muscle/body ache, loss of taste or smell, shortness of breath, and headache were removed from the analysis due to $n \leq 5$ . In the primary school-aged HRE cohort, muscle/body ache, loss of taste or smell, and shortness of breath were removed. In the secondary school-aged HRE cohort, diarrhea, muscle/body ache, shortness of breath, and sneezing were removed.

**FIGURE 1:**
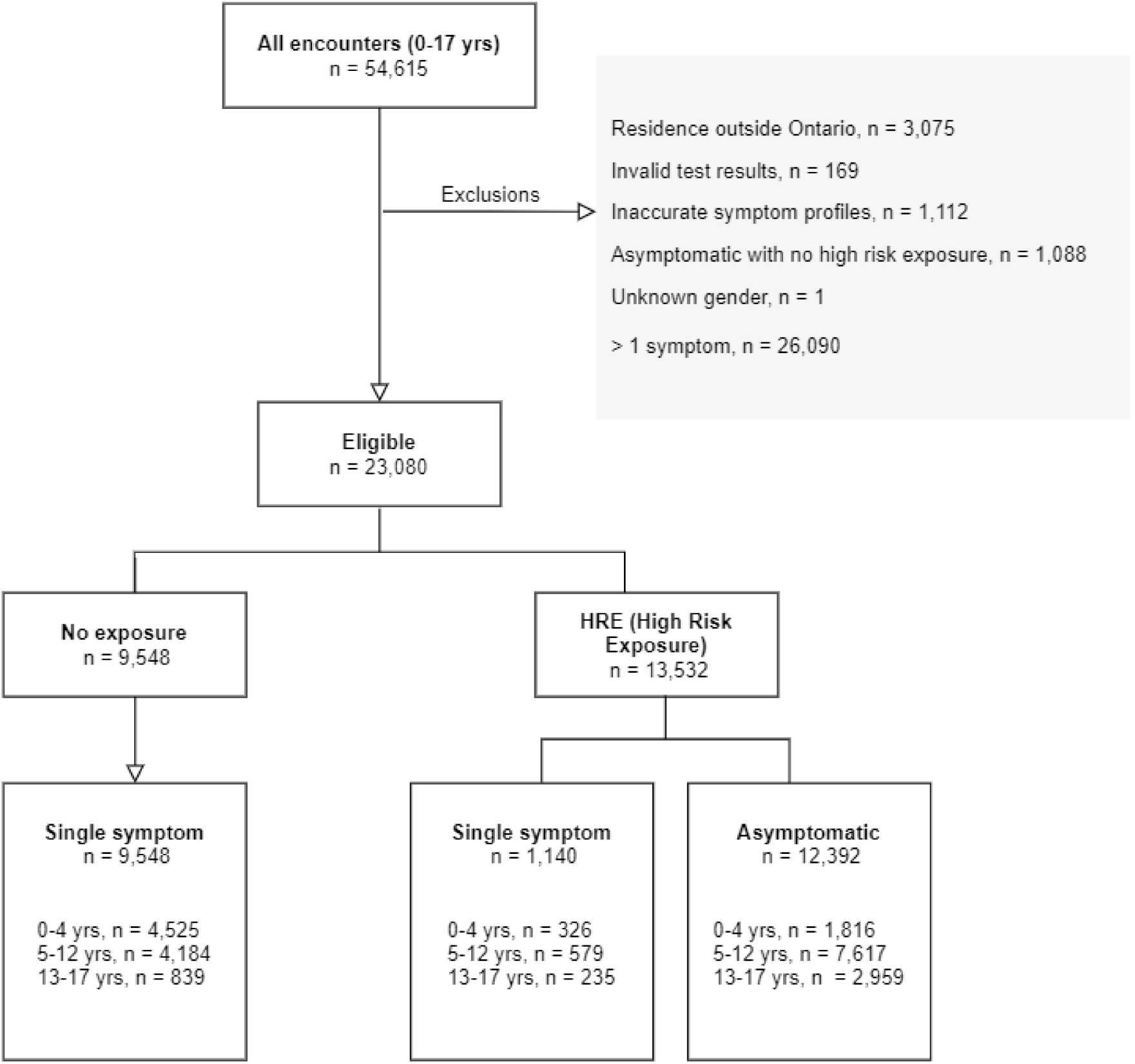
Study flow diagram.

Of the 23,080 pediatric encounters who met inclusion criteria for the study, single symptoms were reported in 46.3% (n=10,688) (Fig 1). The positivity rate was 2.7% (289/10,688), 17.3% (197/1,140), and 5.5% (680/12,392) for all single symptom encounters, HRE single symptom encounters and HRE asymptomatic encounters respectively. Across all multi-symptom patterns, cough and/or rhinorrhea/congestion comprised 35% of all symptomatic presentations (Supplemental Fig 2). As single symptom presentations, rhinorrhea/congestion, cough, fever and sore throat/difficulty swallowing comprised 30% of all encounters, 76% of those involving only a single symptom, and 18.8% of all test-positive cases (Fig 2; Table 1). The relative frequency of single symptoms varied across age groups; in the daycare group, rhinorrhea was the most common single-symptom presentation, followed by fever and cough (43.2%, 23.2%, 17.9%, respectively; Fig 3). Among the primary school age group, rhinorrhea was again the most common presentation, followed by sore throat and cough (24.3%, 19.5%, 17.3%, respectively), whereas the most frequent single symptom in the secondary school age group was sore throat, followed by cough and headache (29.8%, 16.0%, 14.3%).

**FIGURE 2:**
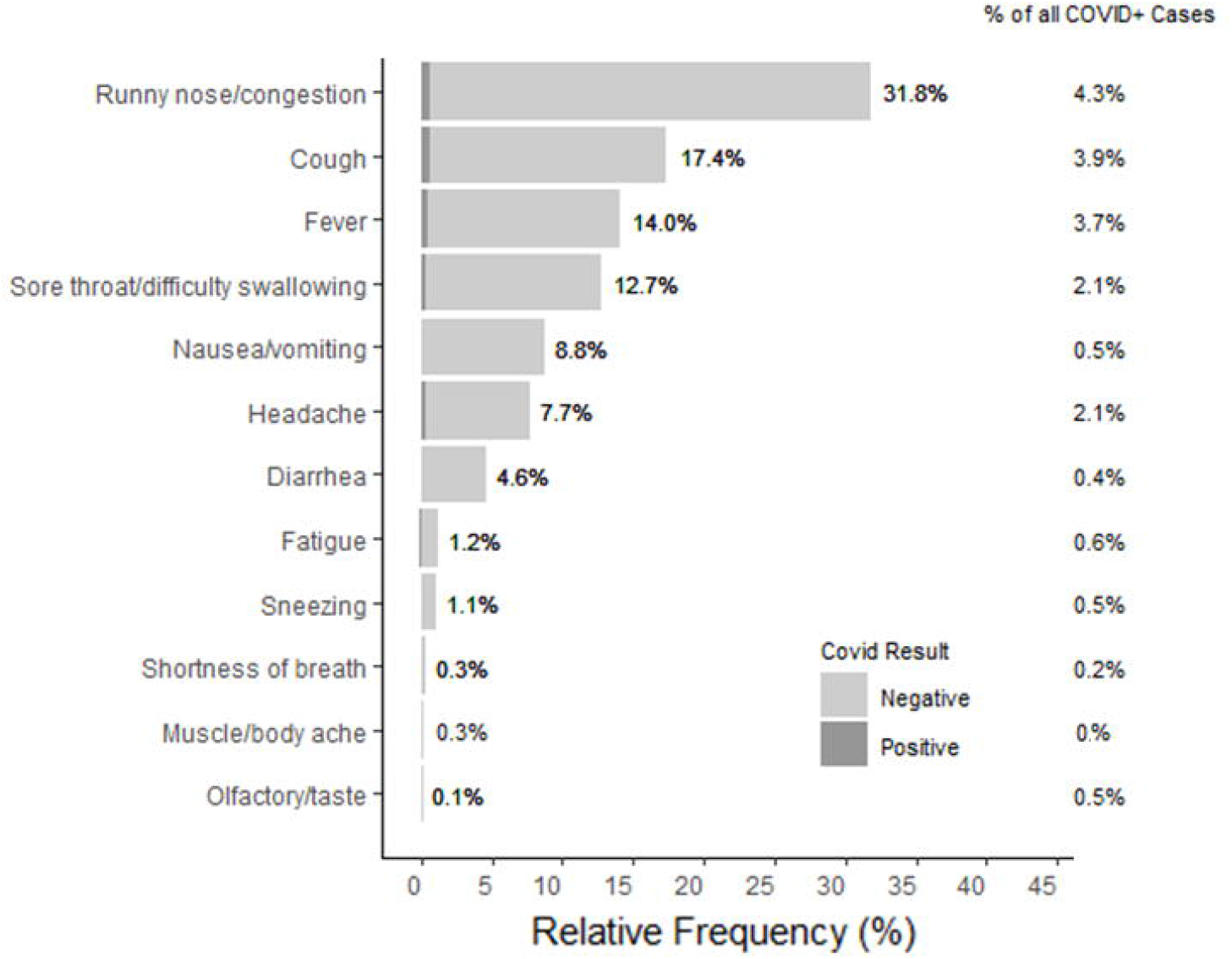
Relative frequencies of single symptoms for all pediatric encounters at a testing centre, shaded by test result as negative (light grey) and positive (dark grey). The right column denotes how positive cases from each single symptom contributed to the total of all COVID-19 positive cases (including single and multi-symptom encounters).

**FIGURE 3:**
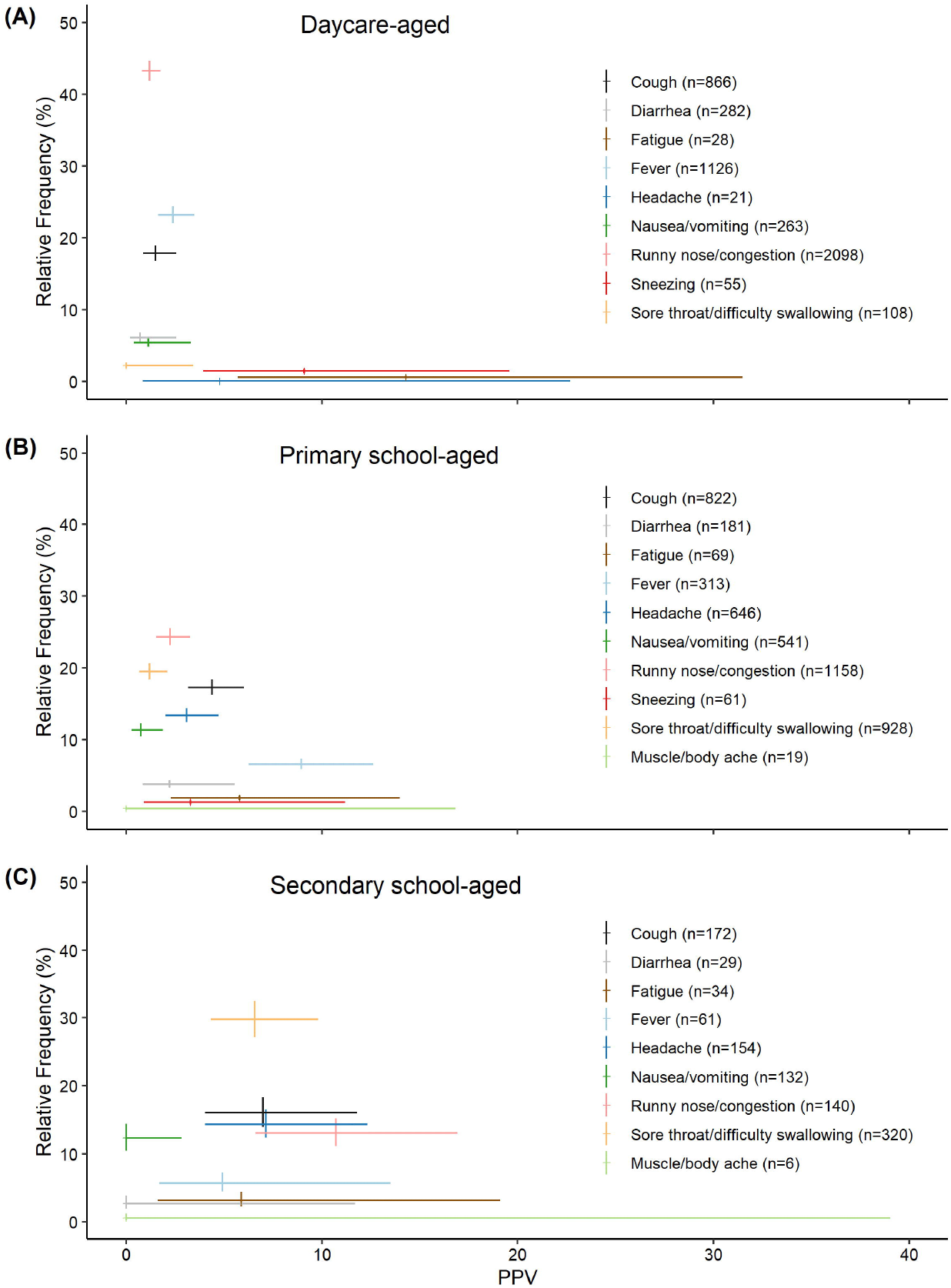
Scatterplot with 95% CI for positive predictive value (PPV) and relative frequency of single symptoms of all comers for (A) daycare, (B) primary school and (C) secondary school age groups. The further right on the x-axis, the more predictive the single symptom. The further up on the y-axis, the more common that single symptom presentation at the assessment centre.

### Positive predictive value (PPV)

Among the 10,688 encounters with single symptoms, test positivity was 17.3% (197/1140) and 1.0% (92/9548) among those with HRE compared to non-HRE, respectively. The PPV for single symptoms was highly variable across age groups (Table 2). PPV for single symptoms was higher in the HRE subset compared to all-comers. We tested if PPV for single symptom HRE was different from the benchmark asymptomatic HRE subgroup. No difference was found for nausea/vomiting, diarrhea, fatigue, and sneezing across all age groups where there were sufficient numbers for analysis. The remaining single-symptom HREs had a higher PPV compared to asymptomatic HRE, but the strength of this effect varied by age group. Rhinorrhea and cough as single symptoms had an elevated PPV among primary school-aged children relative to their asymptomatic peers (*p*<0.001 for each), but this difference disappeared for the daycare age-group (p=0.999 for each). Conversely, presentations with fever alone had an elevated PPV for children under 13 years old relative to their asymptomatic peers (daycare: *p*<0.001; primary school: *p*<0.001), but this difference disappears for the secondary school-aged children (p=0.999). The highest PPV was found in secondary school-aged children who presented with loss of taste/smell (all comers single cohort: PPV=0.67, 95%CI 0.35-0.88; HRE cohort: PPV=0.83, 95% CI 0.44-0.97). Fig 4 shows PPV by relative frequency for each single symptom, stratified by age groups.

**Table 2.** PPV for all comers and HRE cohorts, across age groups. Performance metrics for HRE cohort use asymptomatic HRE as the comparison group.

| Symptom | Age group | All Comers | HRE only |  |  |
| --- | --- | --- | --- | --- | --- |
|  |  | PPV (%) (95% CI) | PPV (%) (95% CI)* | NNT<sup>§</sup> | Likelihood Ratio<sup>+</sup> |
| Runny nose/congestion | 0-4 yrs | 1.2 (0.8, 1.8) | 9.6 (5.3, 15.6); p=0.999 | (19, +Inf.; -21, -Inf.) | 1.0 (0.5, 1.7) |
| Runny nose/congestion | 5-12 yrs | 2.2 (1.5, 3.3) | **11.6 (7.4, 17.0); p<0.001** | 15 (9, 48) | **2.5 (1.5, 3.6)** |
| Runny nose/congestion | 13-17 yrs | 10.7 (6.6, 16.9) | **22.7 (11.5, 37.8); p<0.001** | 6 (3, 18) | **5.7 (2.4, 10.8)** |
| Cough | 0-4 yrs | 1.5 (0.9, 2.6) | 11.4 (5.1, 21.3); p=0.999 | (10, +Inf.; -18, -Inf.) | 1.2 (0.5, 2.4) |
| Cough | 5-12 yrs | 4.4 (3.2, 6.0) | **23.9 (16.5, 32.7); p<0.001** | 5 (4, 9) | **5.7 (3.6, 8.5)** |
| Cough | 13-17 yrs | 7.0 (4.0, 11.8) | **18.2 (8.2, 32.7); p<0.001** | 7 (4, 47) | **4.4 (1.7, 8.7)** |
| Fever | 0-4 yrs | 2.4 (1.7, 3.5) | **36.4 (22.4, 52.2); p<0.001** | 4 (2, 8) | **5.2 (2.6, 9.3)** |
| Fever | 5-12 yrs | 8.9 (6.3, 12.6) | **38.0 (24.7, 52.8); p<0.001** | 3 (2, 5) | **11.3 (6.1, 19.8)** |
| Fever | 13-17 yrs | 4.9 (1.7, 13.5) | 14.3 (0.4, 57.9); p=0.999 | (3, Inf.; -6, -Inf.) | **3.4 (1.5, 10.4)** |
| Sore Throat/Difficulty Swallowing | 0-4 yrs | 0.0 (0.0, 3.4) | 0.0 (0.0, 41.0); p=0.999 | - | - |
| Sore Throat/Difficulty Swallowing | 5-12 yrs | 1.2 (0.7, 2.1) | 9.8 (4.3, 18.3); p=0.40 | (9, +Inf.; -63, -Inf.) | 2.1 (0.8, 3.8) |
| Sore Throat/Difficulty Swallowing | 13-17 yrs | 6.6 (4.3, 9.8) | **20.0 (11.1, 31.8); p<0.001** | 7 (4, 18) | **4.8 (2.4, 8.2)** |
| Nausea/vomiting | 0-4 yrs | 1.1 (0.4, 3.3) | 33.3 (7.5, 70.1); p=0.35 | (2, +Inf.; -15, -Inf.) | **4.8 (1.2, 22.6)** |
| Nausea/vomiting | 5-12 yrs | 0.7 (0.3, 1.9) | 6.7 (0.2, 31.9); p=0.999 | (7, +Inf.; -9, -Inf.) | 1.4 (0.0, 6.1) |
| Nausea/vomiting | 13-17 yrs | 0.0 (0.0, 2.8) | 0.0 (0.0, 41.0); p=0.999 | -22 (-26, -19) | - |
| Headache | 0-4 yrs | 4.8 (0.8, 22.7) | n < 6 | n < 6 | n < 6 |
| Headache | 5-12 yrs | 3.1 (2.0, 4.7) | **21.0 (11.7, 33.2); p<0.001** | 6 (4, 17) | **5.0 (2.4, 8.8)** |
| Headache | 13-17 yrs | 7.1 (4.0, 12.3) | **23.1 (11.1, 39.3); p<0.001** | 5 (3, 19) | **5.9 (2.4, 11.6)** |
| Diarrhea | 0-4 yrs | 0.7 (0.2, 2.5) | 10.0 (1.2, 31.7); p=0.999 | (7, +Inf.; -8, -Inf.) | 1.1 (0.0, 3.4) |
| Diarrhea | 5-12 yrs | 2.2 (0.9, 5.5) | 5.9 (0.1, 28.7); p=0.999 | (8, +Inf.; -10, -Inf.) | 1.2 (0.0, 5.2) |
| Diarrhea | 13-17 yrs | 0.0 (0.0, 11.7) | n < 6 | n < 6 | n < 6 |
| Fatigue | 0-4 yrs | 14.3 (5.7, | 37.5 (8.5, 75.5); p=0.24 | (2, +Inf.; -19, - | **5.8 (1.2, 25.4)** |
|  |  | 31.5) |  | Inf.) |  |
| Fatigue | 5-12 yrs | 5.8 (2.3, 14.0) | 16.7 (2.1, 48.4); p=0.90 | (3, +Inf.; -11, -Inf.) | 3.9 (0.4, 14.3) |
| Fatigue | 13-17 yrs | 5.9 (1.6, 19.1) | 9.1 (0.2, 41.3); p=0.999 | (5, Inf.; -8, -Inf.) | 2.1 (0.4, 7.7) |
| Sneezing | 0-4 yrs | 9.1 (3.9, 19.6) | 22.2 (6.4, 47.6); p=0.72 | (3, +Inf.; -16, -Inf.) | 2.7 (0.5, 7.5) |
| Sneezing | 5-12 yrs | 3.3 (0.9, 11.2) | 7.4 (0.9, 24.3); p=0.999 | (8, +Inf.; -14, -Inf.) | 1.5 (0.0, 4.9) |
| Sneezing | 13-17 yrs | n < 6 | n < 6 | n < 6 | n < 6 |
| Muscle/body ache | 0-4 yrs | n < 6 | n < 6 | n < 6 | n < 6 |
| Muscle/body ache | 5-12 yrs | 0.0 (0.0, 16.8) | n < 6 | n < 6 | n < 6 |
| Muscle/body ache | 13-17 yrs | 0.0 (0.0, 39.0) | n < 6 | n < 6 | n < 6 |
| Shortness of breath | 0-4 yrs | n < 6 | n < 6 | n < 6 | n < 6 |
| Shortness of breath | 5-12 yrs | 5.6 (1.0, 25.8) | n < 6 | n < 6 | n < 6 |
| Shortness of breath | 13-17 yrs | 16.7 (4.7, 44.8) | n < 6 | n < 6 | n < 6 |
| Olfactory/taste | 0-4 yrs | n < 6 | n < 6 | n < 6 | n < 6 |
| Olfactory/taste | 5-12 yrs | n < 6 | n < 6 | n < 6 | n < 6 |
| Olfactory/taste | 13-17 yrs | 66.7 (35.4, 87.9) | **83.3 (35.9, 99.6); p<0.001** | 1 (1, 2) | **99.4 (18.4, Inf)** |

\* P-values<0.05 from two proportion Z-test, with Holm's false-discovery correction, are in bold.
§ Given that $NNT = 1/(\text{Absolute Risk Ratio})$ , where the Absolute Risk Ratio approached zero, negative estimates and +/- infinite were introduced to NNT 95% CI.<sup>29</sup>
+ Bootstrapped likelihood ratios calculated using bootLR::BayesianLR.test.<sup>21</sup>

**FIGURE 4:**
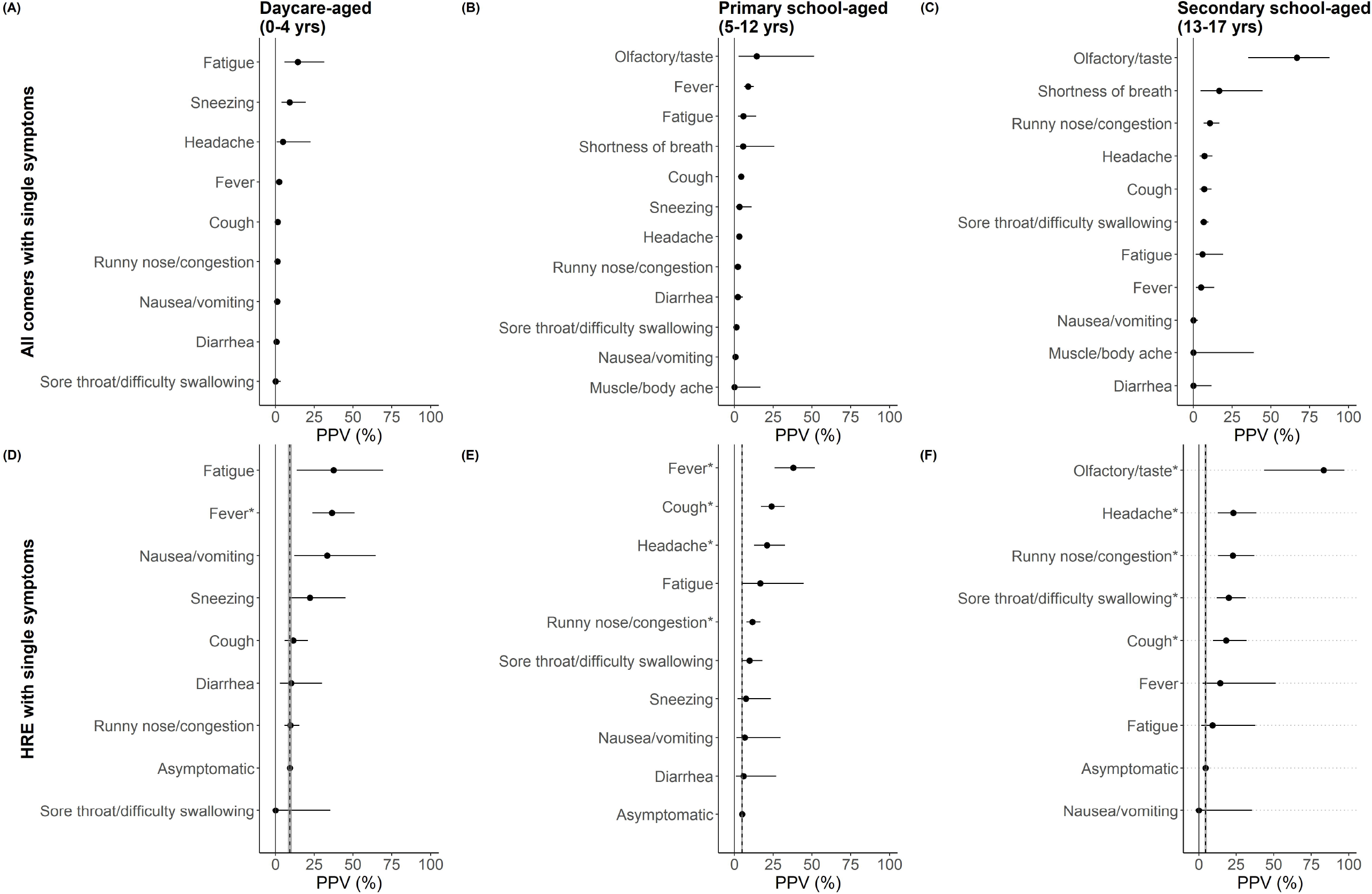
Forest plot showing the positive predictive value (PPV) of single symptoms for all comers (top panel) and among high-risk exposures (HRE) (bottom panel), stratified by age groups. Baseline PPV is shown for the asymptomatic HRE (vertical dashed line=point estimate; grey band=95% CI). Symptoms are starred where PPV is statistically elevated relative to asymptomatic HRE.

### Likelihood ratio

These LRs represent how much more likely is a child with a HRE to receive a SARS-CoV-2 positive test with one specific symptom compared to an exposed child without symptoms (Table 2; Fig 5). Across all age groups, fever as a single presenting symptom was found to increase the probability of SARS-CoV-2 detection. In children aged 0-4 years, the presence of rhinorrhea/congestion or cough had a low LR of 0.9 (95% CI: 0.5-1.5) and 1.2 (95% CI: 0.5, 2.4) respectively. In primary and secondary school-aged children, the presence of rhinorrhea/congestion, cough, sore throat, or headache were each found to increase the probability of SARS-CoV-2 detection, whereas fatigue and nausea/vomiting were more likely to be associated with a positive test in daycare age groups. Consistent between PPV and LR is that loss of taste or smell in secondary school aged children has the highest LR estimate (LR=99.4, 95% CI: 18.6-Inf; omitted from Fig 5). Sneezing and diarrhea did not affect the probability of testing positive for SARS-CoV-2.

**FIGURE 5:**
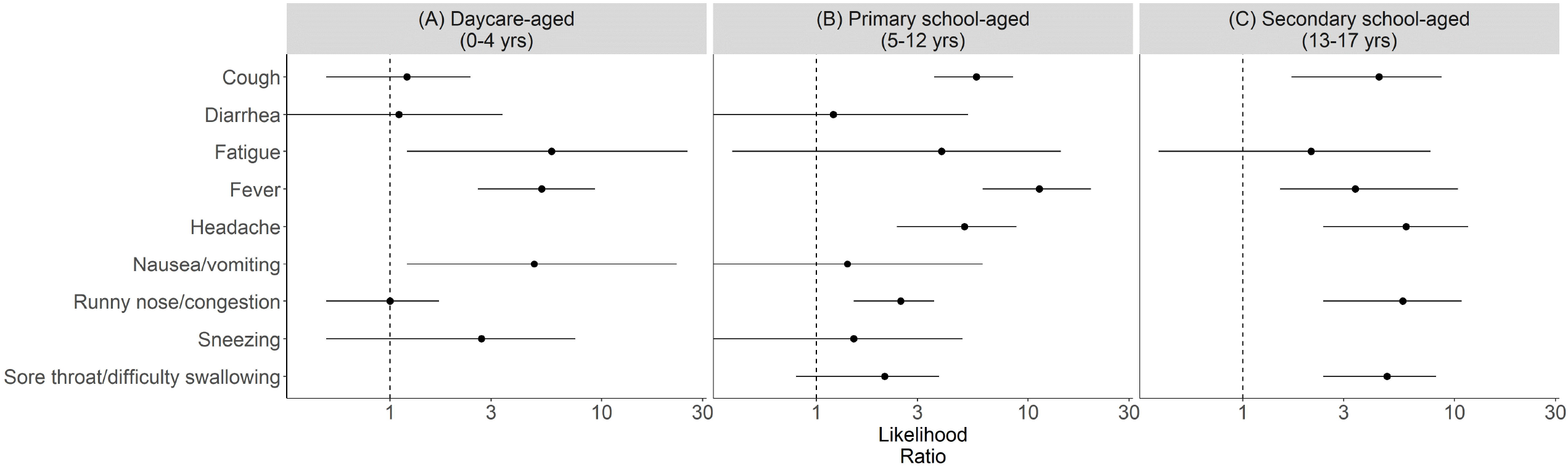
Forest plot showing the positive likelihood ratio for the high-risk exposure (HRE) cohort across (A) daycare, (B) primary school and (C) secondary school age groups. See Table S3 for summary statistics. Where n<6, values were omitted.

### Number needed to test

We sought to determine the number needed to test (NNT) to find one additional SARS-CoV-2-positive case among individuals with HRE presenting with single symptoms, stratified by age groups (Table 2). Among students presenting with isolated fever, 4 daycare-aged (95% CI: 2, 8) and 3 primary school-aged children (95% CI: 2, 5) would need to be tested in order to find one positive case. Among secondary school-aged HRE children with isolated fever, NNT 95% CI had negative estimates, indicating uncertainty about its predictive effect (Table 2).

For isolated cough, the NNT to find one additional positive case among HREs in primary and secondary school-aged groups was 5 (95% CI: 4, 9) and 7 (95% CI: 4, 47), respectively. Rhinorrhea/congestion single symptom screening required 15 primary school-aged (95% CI: 9, 48) and 6 secondary school-aged children (95% CI: 3, 15) to be tested in order to find one positive case; among daycare-aged HRE children with either of these single symptoms, NNT 95% CI had negative estimates, indicating uncertainty about its predictive effect.

## Discussion

In this study at a large urban pediatric COVID-19 assessment centre, 32% of children and youth presented with a single symptom, most commonly rhinorrhea/congestion, followed by cough or fever, and least commonly shortness of breath, muscle/body ache and loss of taste or smell. Fever, as an isolated symptom, increased the likelihood of a positive SARS-CoV-2 test across all pediatric age groups. Yet, the predictive value of rhinorrhea/congestion, the most frequent single symptom presentation in children 12 years and under, was less clear; in children under 5 years, it was not predictive of a positive test even following HRE.

In the daycare age group, combined, rhinorrhea/congestion or cough as a single symptom accounted for 61% of all single-symptom presentations, consistent with provincial patterns.^16^ While rhinorrhea/congestion was not predictive of testing positive for SARS-CoV-2 in the daycare group, the likelihood of a positive test in older ages was elevated.

Our data are consistent with the pediatric literature and regional patterns of multi-symptom presentations.^15, 16^ A particular strength of the study was the stratification by childcare and school age groups, as the predictive values were found to vary accordingly, and can be used to inform screening programs in schools and indoor settings. The goals of any screening program need to be articulated up-front, as there are trade-offs between capturing infrequent presentations with some predictive value, and frequent presentations with high predictive value, depending on the age group. Furthermore, there are opportunity costs to consider with respect to testing, tracing, rate of spread with the presence of variants in the community, and caregiver productivity/exclusion from the workplace. It may be possible to reduce the burden of symptom-based screening tools in schools and childcare settings by including symptoms for the various age groups based on their predictive value. While this may ease the childcare burden on families and testing burden on assessment centres, it should be noted that even less predictive symptoms may represent a significant proportion of missed cases in a community, if the symptom presents frequently. As an example, given the low predictive value of single symptom rhinorrhea/congestion in children aged 0-4 years, this symptom may be considered for removal from daycare screening tools, yet this presentation is noted in nearly 7% of all positive tests in this age group.

During the study period, among children and youth who presented with a single symptom, 2.7 % (n=290) tested positive, yet comprised 19% of all test-positive cases, confirming that there may be utility for single symptom-based screening of SARS-CoV-2 infections, depending on screening program objectives. Some single symptoms, such as diarrhea, have low predictive value and represent a small portion of COVID-19 cases identified during the study period (0.4%). Such symptoms could be considered for removal from criteria for SARS-CoV-2 testing, but remain as criteria for school exclusion since they may reflect other communicable diseases.

The appropriateness of removing a less predictive single symptom from screening tools will depend on screening program objectives within a jurisdiction. Screening programs that aim to identify and exclude all individuals with COVID-19 will include rhinorrhea/congestion as a single symptom. If those with isolated rhinorrhea/congestion during the study period were not screened out and directed to test, 1.6% of identified COVID-19 cases among children and youth would not have been identified. Alternatively, if screening programs aim to identify as many COVID-19 infections as possible with finite resources (but not *all* infections), it may be appropriate to remove less predictive symptoms from screening tools to minimise opportunity costs. For symptoms removed from screening tools, the predictive value among HREs should be considered when determining how to manage those with known exposures. Critically, no research study can determine a threshold appropriate for all jurisdictions, as this will be influenced by local transmission, vaccination rates, and screening program goals and constraints.

This study has several notable limitations. It was undertaken during a period of reduced co-circulation of respiratory viruses due to masking, distancing and stay-at-home orders. As the frequency of non-COVID-19 respiratory viruses and their associated symptoms increase, the our results may overestimate the utility of single symptom screening when masking and social distancing measures diminish. Another feature of importance is the low prevalence of variants of concern during most of our study period. If emerging variants shift their symptom profiles, the generalisability of these findings to new variants should be revisited. ^9, 23^

Symptoms and HRE status were self-reported, therefore children/parents may not report symptoms that are mild or incur misreporting. Some parents may have experienced a greater financial burden of isolation for a pending or positive result,^24, 25^ which may produce an inherent socioeconomic bias to the under-reporting. As well, from Oct 1-Feb 21, non-HRE people with mild single symptoms were not directed for testing unless the symptom persisted >24hrs and/or new symptoms emerged. However, earlier return to school, daycare, or the workplace (for the affected adults in the household who needed to isolate pending the result of their child’s test) was possible with a negative test and symptom resolution. As such, there was no change in the time to get tested between periods when schools were open or closed to in-person learning.

The number needed to test provides insight into the consequences associated with a testing strategy. While there are limitations with the NNT metric ^26^, its utility was considered for population-based recommendations, acknowledging the number of children whose learning would be disrupted for every positive case detected.

Our two-cohort approach aims to minimize risk of selection bias such as Berkson’s paradox. Our concern was that those children and youth arriving at testing centres were different from the general population (e.g., presenting with more severe symptoms, risk sensitivities, more health literate, socio-economic makeup). This bias is called Berkson’s paradox, and has been a noted limitation for studies in which data collection is focused on assessment centres ^27, 28^. We assume that the symptomatic children in the HRE cohort made the decision to get tested primarily due to an exposure (rather than symptom severity), while single-symptom cohort encounters may be reliant on perceived severity of symptoms by parents or daycare/school staff. Therefore, within the HRE cohort, the range of single-symptom severity should more closely reflect that of the general population.

Decisions around screening and testing for SARS-CoV-2 aim to strike a balance between finding all cases and minimizing learning/childcare disruption. When measures are in place within schools to prevent transmission, including masking, cohorting and/or distancing, in-school transmission has been shown to be low, suggesting a more targeted screening program would still be effective.^8^ As measures are relaxed in the schools, transmission may be more significant, especially in the vaccine-ineligible group of children under age 12 years, and with a more contagious strain. At such time, case-finding with broader screening criteria will be more important.^6, 14^ Regardless of screening strategy, robust contact tracing and low-barrier testing strategies will be required to ensure that every patient who screens positive can access easier and faster testing. Where the prevalence of circulating SARS-CoV-2 is low and contact tracing resources are preserved, individuals with infectious symptoms should continue to self-isolate during the period of their illness, however the criteria for SARS-CoV-2 testing may be refined to the more predictive symptoms to improve testing uptake, isolation adherence and minimization of learning disruptions in communities.

## Conclusions

Educational settings during the pandemic have faced trade-offs between in-person learning and exclusion of those with symptoms and exposures to prevent introduction of SARS-CoV-2 into daycare and school settings. This study shows that single symptom presentations contribute a substantial proportion of testing, and provides predictive value based on the pediatric age group. Single symptoms that predicted SARS-CoV-2 infection were; fever across all age groups; nausea/vomiting for the under 5 year olds, headache, rhinorrhea/congestion and cough for school aged children and youth; and sore throat for secondary school aged youth. Neither rhinorrhea/congestion nor cough predicted SARS-CoV-2 infection in those under 5 years, despite contributing a high testing volume. Decisions around symptom-based screening should balance emerging evidence, while considering the goals of the screening program and local context.

## Data Availability

Data are available upon request

## Abbreviations

(SARS-CoV-2): Severe acute respiratory syndrome coronavirus 2
(COVID-19): Coronavirus disease 2019
(HRE): High-Risk Exposures
(PPV): Positive Predictive Value
(LR): Likelihood Ratio
(NNT): Number Needed to Test

## Acknowledgments

Nick Barrowman and others at the Clinical Research Unit, CHEO Research Institute, provided epidemiological and statistical comments. We would also like to thank all the frontline assessment centre workers who collected these data.

## Captions for figures

**SUPPLEMENTAL FIGURE 1:** SARS-CoV-2 Rates/100,000 for Ottawa residents aged 0 to 17 years from October 1, 2020 – April 25, 2021 (N = 3,172 cases).

**SUPPLEMENTAL FIGURE 2**: Ranked bar showing the relative frequencies of all clinical presentations (including single and multi-symptom patterns; n=36,788) among all children and youth (aged 0 to 17 years). Data from Oct 2020-April 2021.

