## Supplementary figures and images for "Predicting SARS-CoV-2 infections for children and youth with single symptom screening"

### Supp Fig 1

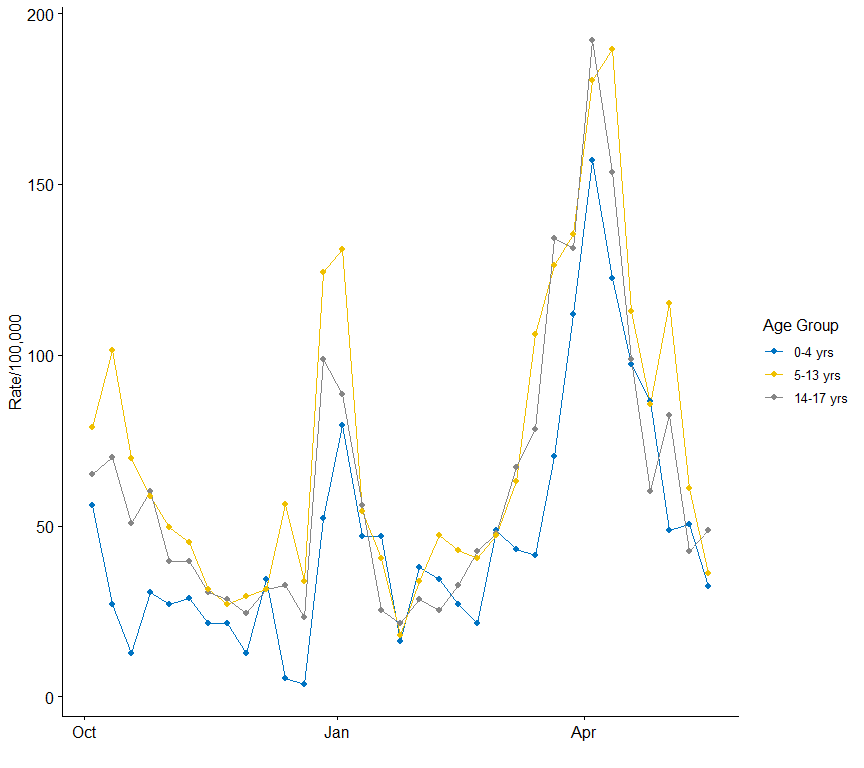

### Supp Fig 2

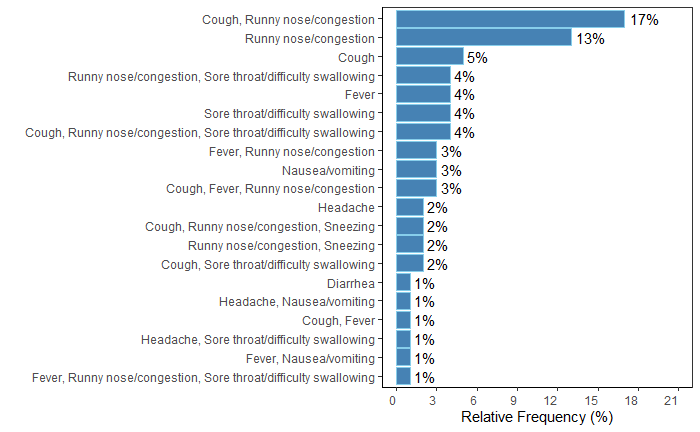
